# Expanding Covid-19 Testing: Mathematical Guidelines for the Optimal Sample Pool Size Given Positive Test Rate

**DOI:** 10.1101/2020.05.21.20108522

**Authors:** Kayleigh Adams

## Abstract

Widespread testing is essential to the mitigation of the spread of any virus, and is particularly central to the discussion on transitioning out of national quarantine. Sample pooling is a method that aims to multiply testing capability by using one testing kit for multiple samples, but will only be successful under certain conditions. This paper gives precise guidelines on those conditions for success: for any proposed sample pool size, explicit bounds on the positive infection rate are given that are informed by both discrete and statistical modeling.

## Sample Pooling to Get More Out of Testing

Sample Pooling is a method of using one test for multiple samples with the aim of using less tests on more people. In Israel, researchers have been testing up to 60 samples at a time for COVID-19 (https://www.hospimedica.com/coronavirus/articles/294781273/israeli-researchers-introduce-pooling-method-for-covid-19-testing-of-over-60-patients-simultaneously.html). The Governor of Nebraska, Pete Ricketts, has just proposed that hospitals in his state test 5 samples at once (https://nebraska.tv/news/local/governor-ricketts-state-working-to-expand-coronavirus-testing).Rather than testing samples individually, *n* samples are placed into the same test tube, a sample pool of size *n*. If the test comes back negative, then all *n* samples are reported negative. If the test comes back positive, then the samples must be re-tested. This method, if used under the right conditions, has the potential to vastly expand testing capacity, meaning we can more readily isolate those that are infected and prevent them from spreading the disease any further. Intuitively, it seems that this method would cause problems if a very large portion of the sample population was infected with the pathogen of interest, or if the number *n* was too large.Through a simple discrete model and a statistical model, we can make explicit this relationship between “efficiency”, the size *n* of the sample pools, and the positive test rate of the sample population, which we will call *p*.

## A Discrete Model for a Guaranteed Upper Bound

If a sample population is split into pools of n samples, then we define *G^−^* and *G*^+^ to be the number of groups or pools of samples to test negative and positive, respectively. The samples in a negative group will only use 1 test, while the samples in a positive group will use *n* + 1 tests due to re-testing. Then we can say that the *n* sample pooling method is *efficient* if it uses less tests than individual testing, that is, if the following is true:

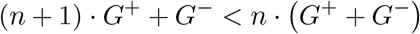

Which reduces to

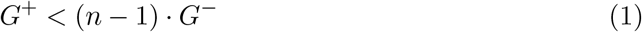

From here, we can find a guaranteed upper bound for the positive test rate as a function of *n* that would guarantee “efficiency”.

We call this upper bound *guaranteed* because it does not find the positive test rate that would *statistically* guarantee efficiency, but that would always guarantee it, even in extremely unlikely scenarios.

The configuration of positive samples that would give the lowest possible positive test rate is having *exactly* one positive sample in every positive group *G*^+^.

Then the formula for the positive test rate of the sample population is the following:

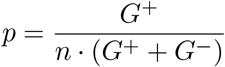

Using inequality (1), we get

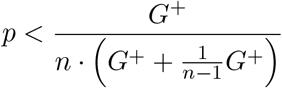

Which simplifies to

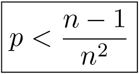

To give this some context, if Nebraska wants to use its proposed method of sample pooling of size 5, they can guarantee that tests will not be overused if the positive test rate of people getting tested is less that 16%.

It is also important to note that this discrete method cannot guarantee that pooling of any size will be more efficient if the positive test rate is above 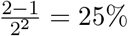.

## Statistical Upper Bounds

Our statistical model is pretty simple, and without loss of generality we set our number of tests available, *t_A_* to be 2000 in our simulations.

To explain this model, we will find how many tests are required to test *S* samples using sample pools of size *n* at positive test rate *p*.

In the case of sample pooling of size *n*, we assume that we have *S*/*n* sample groups. For each positive test rate *p*, the probability of a sample group needing to be re-tested is 1 − (1 − *p)^n^*.

So, we assume that for positive test rate *p*, 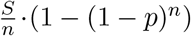 groups need to be re-tested. So, the number of tests required, *t_r_* is then:

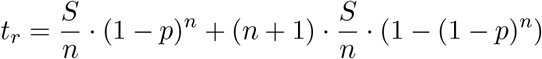

So, 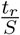 tests are required per sample, and the inverse of that is that 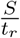 samples can be tested per test.

Then the number of samples we are capable of testing with *t_A_* available tests, positive test rate *p*, and sample pool size *n*, is 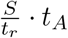.

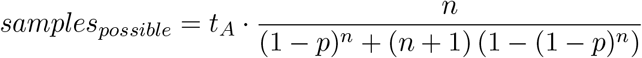

Clearly *t_A_* is a scalar here, which is why our choice of *t_A_* = 2000 does not lose generality.

In the appendix there are some results of this model, including the minimum pool size of 2, Nebraska’s proposed pool size of 5, and Israel’s pool size of 60. The intersection points are highlighted, as their *p* values represent the “statistical upper bound”.

## Conclusion

Both the conservative discrete model and the realistic statistical model suggest that sample pooling can be very beneficial in expanding testing, which is crucial to stemming the spread of any outbreak, particularly Covid-19. The models also show that sample pooling could potentially be a waste of resources, and give precise conditions that the determine the help or harm that sample pooling can present. Sample pooling should be used in areas that are not so severely hit (< 25 − 30%) to prevent a wider spread, and/or conserve resources to be shared with areas that are suffering from higher positive test rates, where sample pooling is absolutely not an option. The positive test rate in the US as of 5/20/2020 according to Johns Hopkins University of Medicine is 12.3%, which is viable for pooling. Since even our conservative model has suggests sample pooling is efficient if the positive test rate is less than 25%, much of the US should consider sample pooling.

It is important to note that testing is not only important when the outbreak is surging, but will also be very important after the peak as we consider relaxing quarantine and social distancing measures. Since sample pooling is more powerful as the positive test rate decreases, we can dramatically expand testing after the cases drop. With the extra testing ability due to sample pooling, we could test people as a prerequisite to going back to work, ensuring the safest possible transition out of quarantine. Samples from all members of a household could be pooled, and need not be retested because if any member of the household is positive, then no member should return to work.

With expanded production of tests and sample pooling, we can expand testing beyond what we thought possible. As the economy re-opens, we will need extensive testing to prevent a resurgence of this disease. Informed by the results of these two models, we can take a very safe, and very efficient, road to recovery.

## Data Availability

This paper is theoretical and does not use data

## Acknowledgements

Thank you to Mary Zhang for introducing me to sample pooling and thank you to both Mary Zhang and Lisa Larsson for discussing this interesting problem with me.

## Appendix

**Figure 1:**
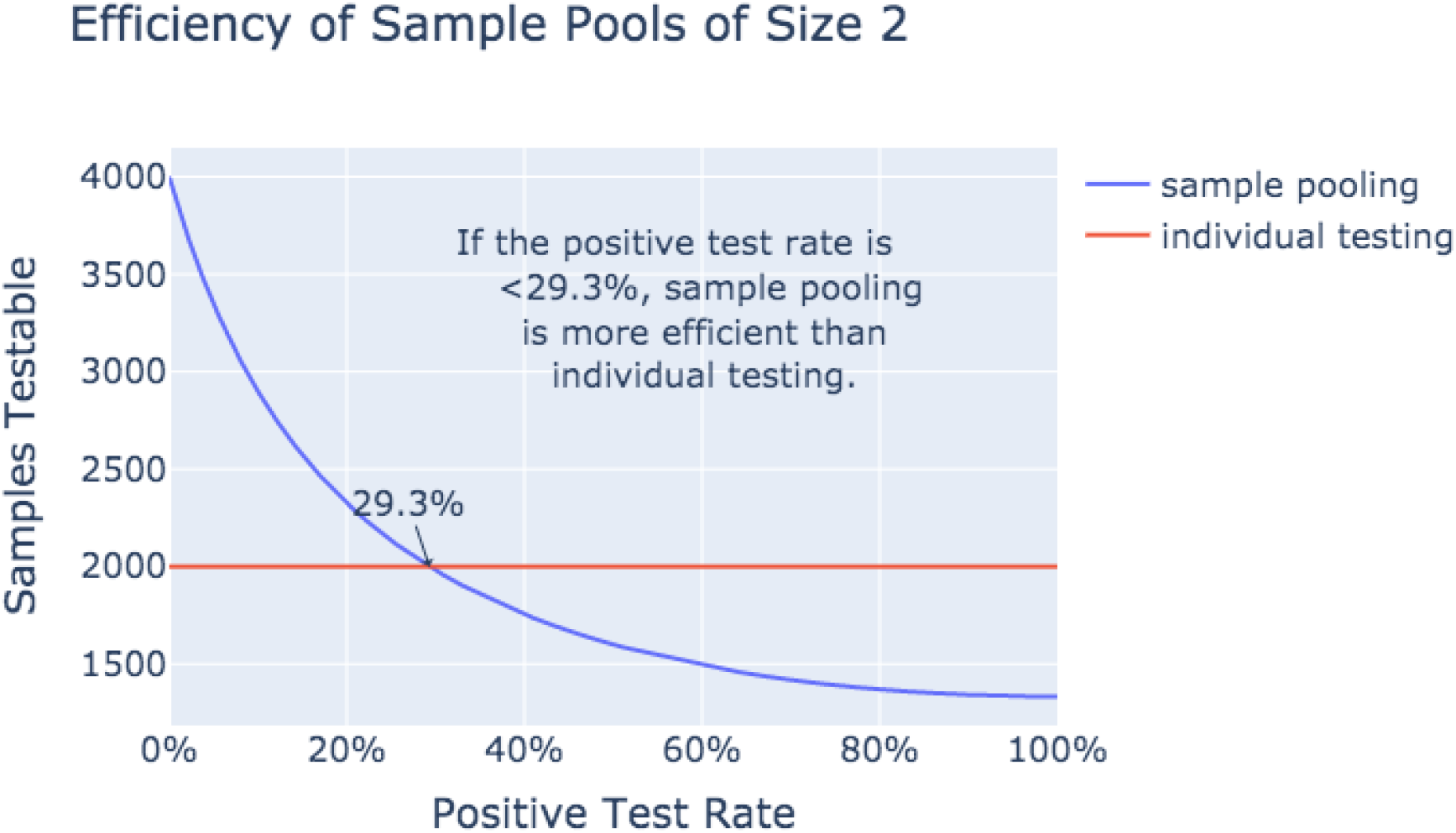
Number of Samples Testable Given 2,000 Tests vs. Positive Test Rate: Sample Pool Size 2, Highlighting that More Samples Can Be Tested *p* < 29.3%

**Figure 2:**
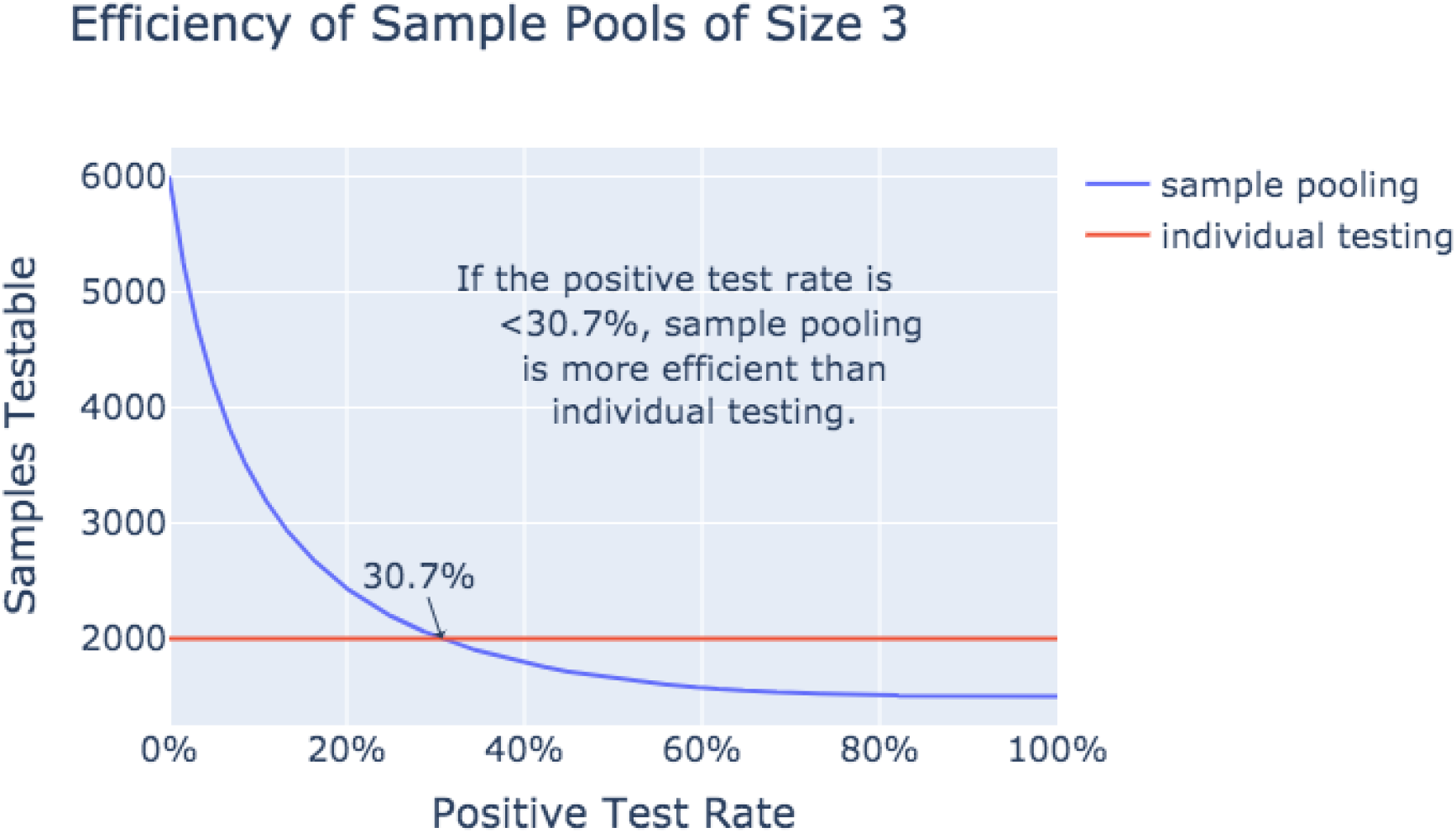
Number of Samples Testable Given 2,000 Tests: Sample Pool Size 3 vs. Positive Test Rate *p*, With Higher Efficiency Than Individual Testing for *p* < 30.7%

**Figure 3:**
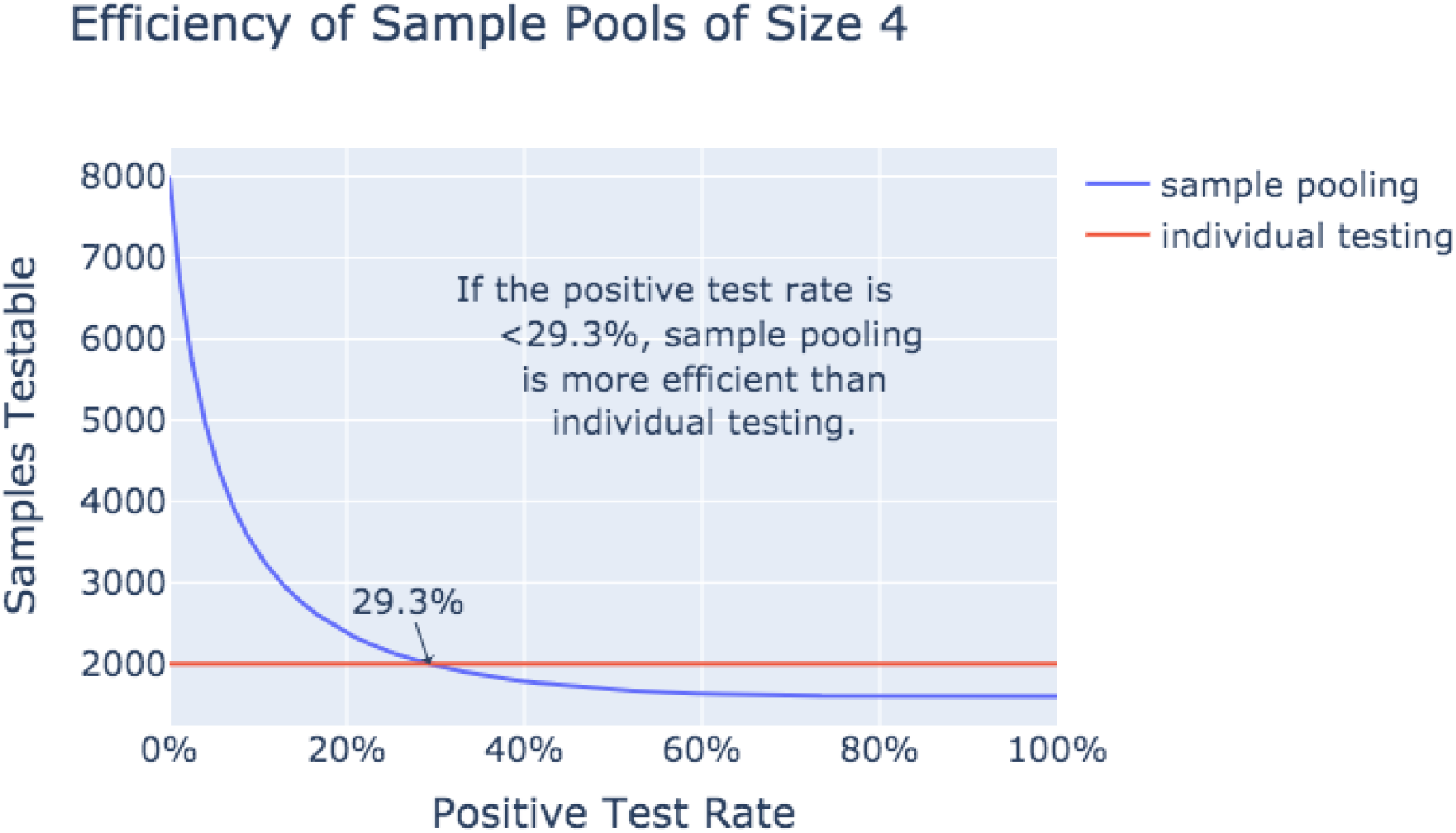
Number of Samples Testable Given 2,000 Tests: Sample Pool Size 4 vs. Positive Test Rate *p*, With Higher Efficiency Than Individual Testing for *p <* 29.3%

**Figure 4:**
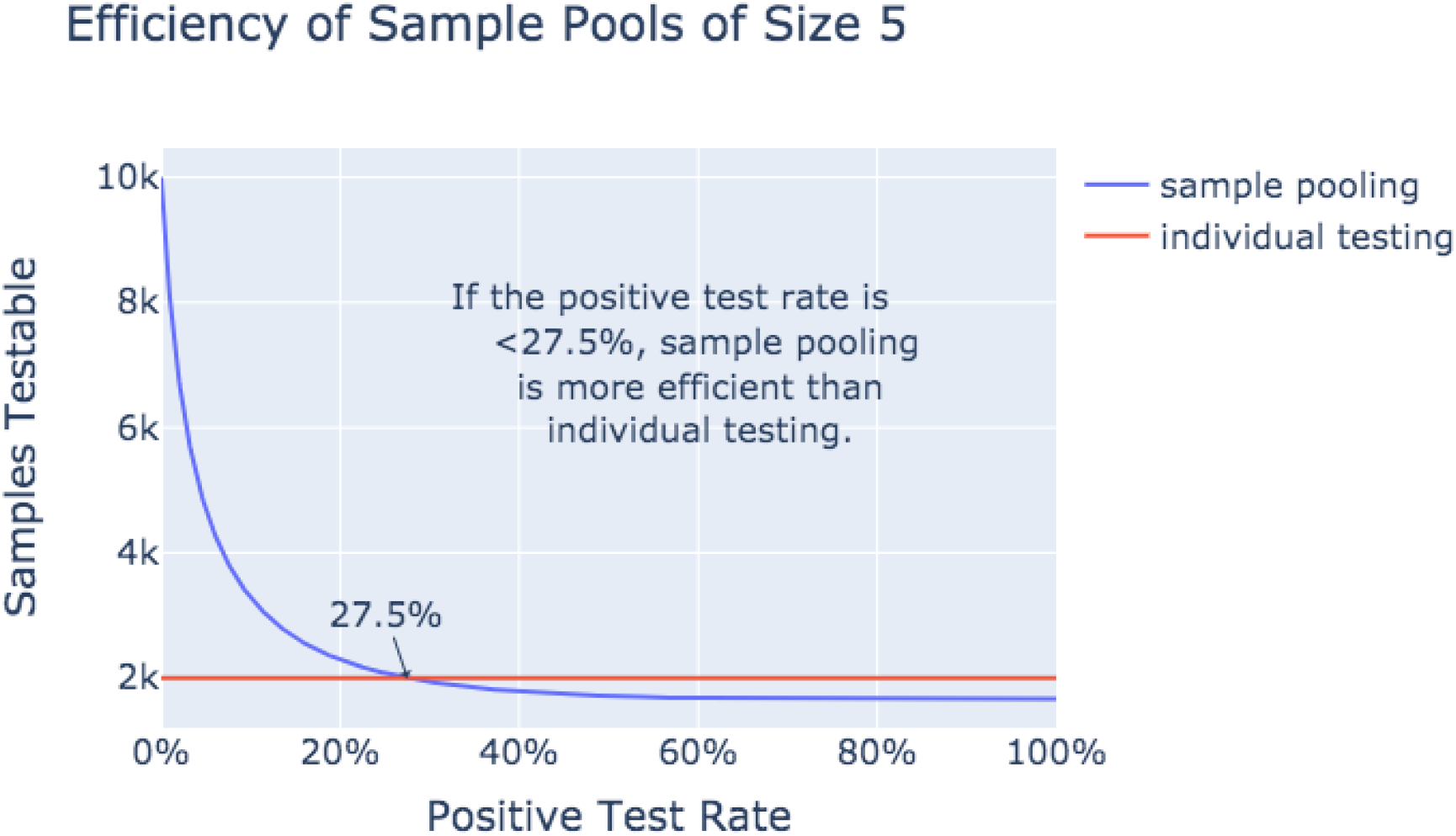
Number of Samples Testable Given 2,000 Tests: Sample Pool Size 5 vs. Positive Test Rate *p*, With Higher Efficiency Than Individual Testing for *p* < 27.5%

**Figure 5:**
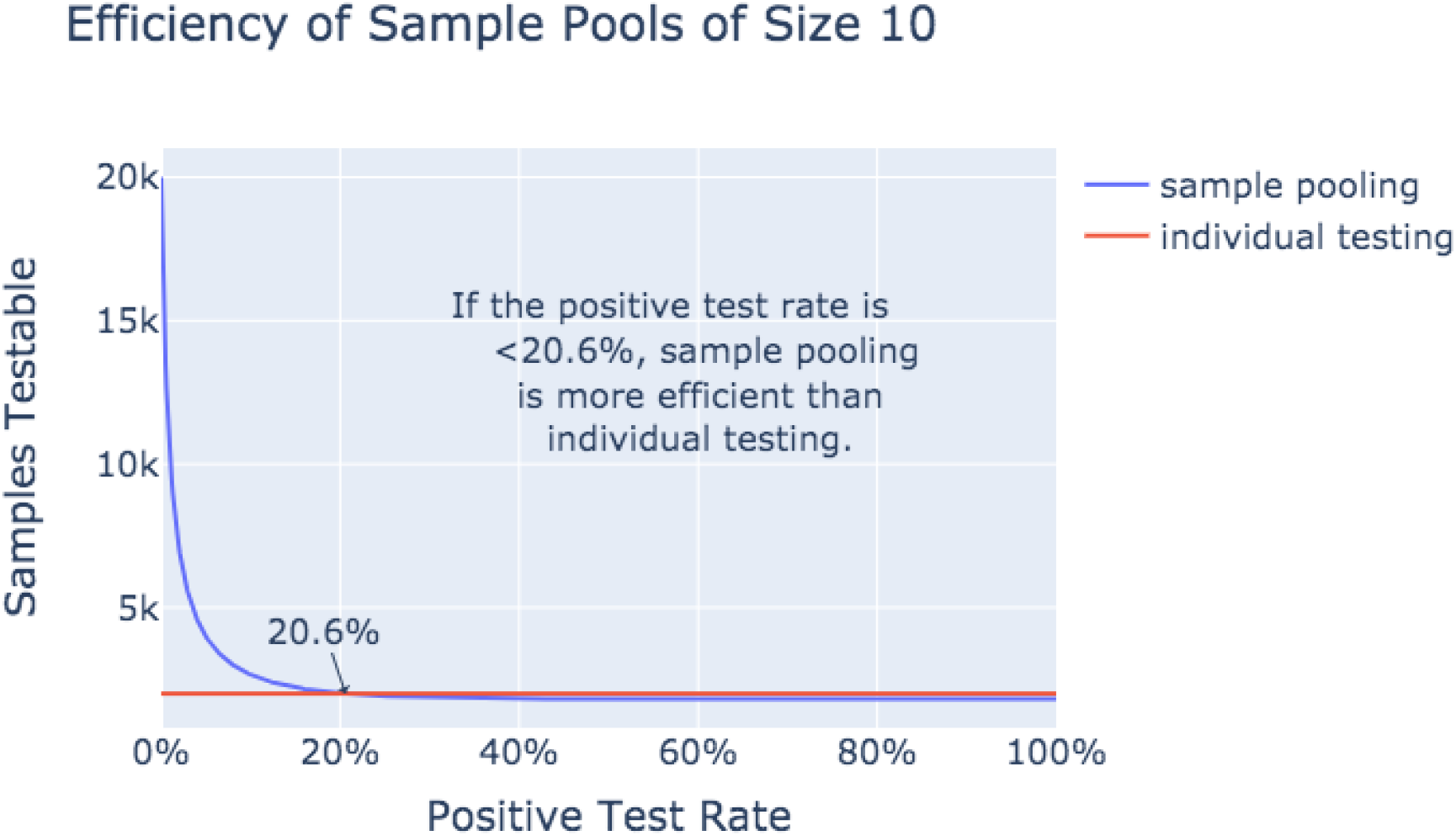
Number of Samples Testable Given 2,000 Tests: Sample Pool Size 10 vs. Positive Test Rate *p*, With Higher Efficiency Than Individual Testing for *p* < 20.6%

**Figure 6:**
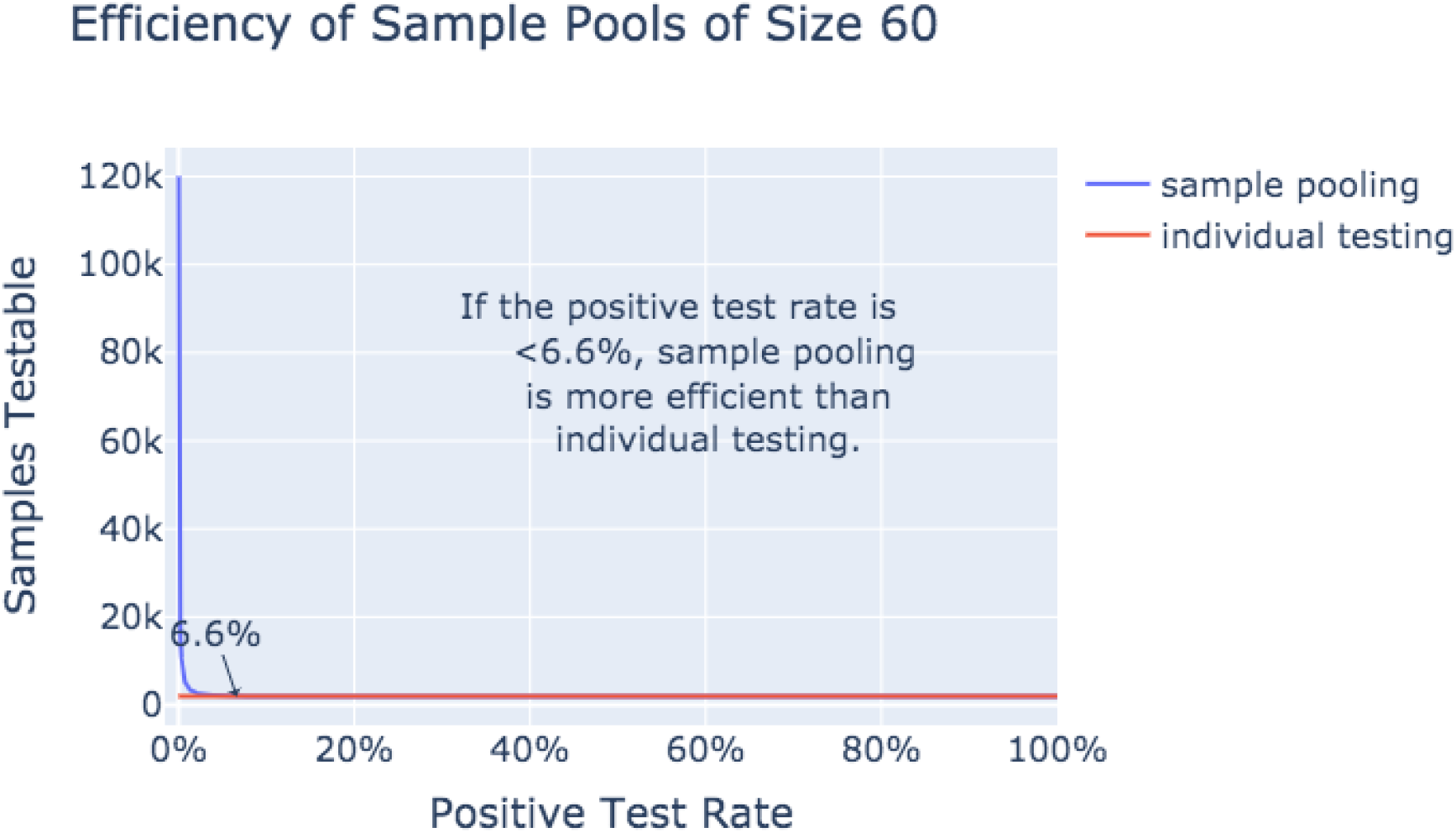
Number of Samples Testable Given 2,000 Tests: Sample Pool Size 60 vs. Positive Test Rate *p*, With Higher Efficiency Than Individual Testing for *p* < 6.6%

